# Multi-unit activity and high-gamma patterns dissociate in the seizure onset zone during focal to bilateral tonic-clonic seizures

**DOI:** 10.1101/2025.11.03.25339320

**Authors:** Urszula Gorska-Klimowska, Beril Mat, Colin Denis, Brinda Sevak, Csaba Kozma, Dillon Scott, Mariel Kalkach Aparicio, Cynthia Papantonatos, Aaron J. Suminski, Maximilian Grobbelaar, Elena Monai, Felipe B. de Paiva, Aaron F. Struck, Giulio Tononi, Wendell Lake, Melanie Boly

## Abstract

**Objectives:** To track multi-unit activity (MUA) using hybrid Behnke–Fried electrodes continuously throughout human seizures, and to characterize how both neuronal firing and high-gamma (HG) changes over the ictal course, within versus outside the seizure-onset zone (SOZ), and by seizure severity.

**Methods:** We analyzed 32 seizures - 9 focal preserved consciousness (FPC), 17 focal impaired consciousness (FIC), 6 focal-to-bilateral tonic–clonic (FBTC) - from 8 patients implanted with Behnke–Fried hybrid depth electrodes. Spikes were sorted jointly across a 10-min pre-ictal baseline and ictal period, yielding 895 units trackable through seizures. Macroelectrode contacts within 15 mm of each microelectrode were classified by the SOZ status of the corresponding micro site. Firing rates from MUA, HG power and phase-locked high-gamma (PLHG) were baseline-normalized and compared across seizure types and periods (first/second half; pre/post-generalization).

**Results:** Unit waveforms were identified throughout seizures. Within the SOZ, firing increased at onset in all types, then attenuated (FPC, FIC) or plateaued (FBTC). PLHG peaked early and declined across all seizure types, including post-generalization in FBTC. In contrast, HG stayed elevated at onset for all seizures, was highest in FBTC, and increased further after generalization. Outside the SOZ, MUA, HG, and PLHG stayed near baseline in FPC, and rose progressively in FIC. In FBTC they showed a temporal dissociation: HG/PLHG rose pre-generalization and remained high, while MUA increased sharply only after generalization.

**Significance:** We demonstrated feasibility to track multi-unit activity throughout seizures and revealed seizure-type– and location-specific micro–macro dynamics. Collectively, early rise and later decline in PLHG that mostly aligns with neuronal firing was concordant within SOZ. Moreover, increases in all measures outside the SOZ are a hallmark of seizures with impaired consciousness, and may suggest presence of a third driver. These findings offer testable biomarkers for future larger clinical studies.

**KEY POINTS:**

- MUA can be tracked continuously during seizures with Behnke–Fried hybrid electrodes.
- In the SOZ, firing rises at onset, then attenuates in FPC/FIC or plateaus in FBTC; HG rises while PLHG peaks early then declines.
- Outside SOZ, firing, and HG/PLHG progressively increase, specifically after generalization.
- Combined MUA/HG/PLHG provide candidate biomarkers for SOZ mapping and impaired consciousness.

## 1. INTRODUCTION

Loss of consciousness (LOC) is a hallmark of many epileptic seizures and is associated with risks of injury, impaired quality of life, and increased mortality (Manford et al., 1996). A clearer mechanistic account of ictal LOC is clinically important because it can help prioritize neuromodulation targets and timing—for example, distinguishing seizure types and network states that may benefit from thalamic stimulation strategies (Englot et al., 2010; Doss et al., 2024; Pizzo et al., 2024; Fisher et al., 2023). Our recent work suggests that LOC arises through distinct physiological mechanisms in focal to bilateral tonic–clonic (FBTC) versus focal impaired consciousness (FIC) seizures (Juan, Górska, Kozma et al., 2023). In FBTC, high-gamma (HG) power in the posterior cortex rises early in the ictal course and tracks behavioral unresponsiveness, and spreads to the front after generalization. In contrast, FIC seizures show widespread increases in slow-wave activity (SWA) as the dominant signature of LOC (increases larger than in FBTC), with more circumscribed HG increases near the seizure-onset zone (SOZ). Because macroelectrodes average activity over millimeters of cortex, large ictal field potentials do not by themselves establish local neuronal recruitment —they can reflect inhibitory processes or distant sources, and alterations in LFPs may occur without increased local firing (Schevon et al., 2012; Merricks et al., 2015). Since spikes are the fundamental output of cortical circuits, microelectrode recordings are required to determine whether LFP signatures of LOC correspond to actual changes in neuronal firing (Weiss et al., 2013; Weiss et al., 2015).

In Utah-array recordings from two patients (three FBTC; Juan, Gorska, Kozma et al., 2023) with arrays placed remote from the SOZ, we found that neuronal firing was heterogeneous before behavioral generalization but rose sharply and remained elevated thereafter. This firing coincided with phase-locked high gamma (PLHG) increases—an index of locally synchronized spiking and ictal recruitment— and with a whole-brain HG peak at generalization, consistent with macro–micro coupling of high-frequency activity in FBTC (Weiss et al., 2013; Smith et al., 2016). However, differences in firing were not systematically assessed for FIC or focal preserved consciousness (FPC) seizures, and firing patterns between SOZ versus surrounding cortex remained untested.

The present study addresses those gaps by examining multi-unit activity (MUA) with Behnke–Fried hybrid depth electrodes during clinical monitoring. Unlike Utah arrays, which sample a high-density cortical patch, Behnke–Fried electrodes allow simultaneous micro- and macro-recordings across broader clinical coverage, including SOZ and surrounding regions (Chari et al., 2020; Guth et al., 2021). This design enables direct comparison of neuronal firing between SOZ and non-SOZ sites across seizure types and links micro-with macro-scale activity. Specifically, we asked: (i) how neuronal firing evolves across ictal periods in FPC, FIC, and FBTC seizures; (ii) whether changes differ between SOZ and non-SOZ contacts; and (iii) how microelectrode firing relates to macroelectrode markers of ictal dynamics, including HG, and PLHG.

## 2. MATERIALS & METHODS

### Datasets

All procedures were approved by the institutional review board for human studies at the University of Wisconsin-Madison, and informed consent for the microelectrode study was obtained from all participants.

Ictal recordings were retrospectively collected from medical records of the University of Wisconsin-Madison (UW-Madison) Hospital and Clinics (UWHC). A total of 32 seizures from 8 patients (6 female, median age 40 years, range 18–57) implanted with Behnke-Fried hybrid depth electrodes (combining macro- and micro-contacts) were included. Microelectrode recordings were acquired using the Blackrock Microsystems system (Blackrock Neurotech, UT) sampling at 30 kHz with 0.3 – 7.5 kHz hardware Butterworth band-pass (1st-order high-pass, 3rd-order low-pass). Macroelectrode recordings were acquired using Natus Quantum system (Natus Medical Inc., Pleasanton, CA) during clinical monitoring at the UW Hospital Epilepsy Monitoring Unit.

The dataset included 9 Focal Preserved Conscious (FPC) seizures, 17 Focal Impaired Consciousness (FIC), and 6 focal to bilateral tonic-clonic (FBTC) seizures. The number of seizures per patient ranged from 1 to 10 (mean: 4, SD: 3.2). Most seizures originated from the limbic lobe (62.5%) and the right hemisphere (54%), and the majority of seizures originated from wakefulness (75%; see Table 1). For each seizure, the timing of onset and offset was determined by a certified epileptologist (MB) using visual analysis. The seizure onset zone was defined based on the initial channels exhibiting epileptiform activity.

**Table 1.** Patient’s clinical characteristics split by seizure type.

|  | FPC | FIC | FBTC | Total |
| --- | --- | --- | --- | --- |
| Patients | 3 | 6 | 5 | 8 |
| Seizures | 10 | 16 | 6 | 32 |
| Seizures per patient, n | 3.33 ± 1.69 | 3.00 ± 1.09 | 1.20 ± 0.40 | 3.87 ± 2.89 |
| Primary seizure focus n (%) |  |  |  |  |
| Limbic | 9 (90) | 9 (56) | 2 (33) | 20 (63) |
| Temporal | 1 (10) | 4 (25) | 2 (33) | 7 (22) |
| Frontal | 0 (0) | 2 (13) | 1 (17) | 3 (9) |
| Frontal and temporal | 0 (0) | 1 (6) | 1 (17) | 2 (6) |
| Seizure arising from sleep, n (%) | 1 (10) | 7 (44) | 0 (0) | 8 (25) |
| SEEG contacts per seizure | 69.2 ± 5.7 | 87.6 ± 17.1 | 84.5 ± 17.2 | 80.0 ± 16.4 |
| Microelectrodes in SOZ/ non-SOZ<br>(8 contacts per microelectrode) | 48/ 120 | 40/ 264 | 24/ 64 | 112/ 448 |
| Macroelectrode contacts in SOZ/ non-SOZ | 76/148 | 50/288 | 34/88 | 160/524 |

Microelectrodes were located within the seizure onset zone (SOZ) in 13 seizures from 4 patients, and outside the SOZ in 30 seizures from 7 patients (number of contacts summarized in Table 1), enabling comparison of firing rates between SOZ and non-SOZ sites.

### Seizure categorization and behavioral scoring

Seizures were classified based on observed behavioral characteristics during the ictal period, following the operational classification of seizures by the International League Against Epilepsy (Beniczky et al., 2025; Fisher et al., 2017). Focal to Bilateral Tonic-Clonic (FBTC) seizures were defined by an initial focal onset followed by generalized symmetric or asymmetric bilateral motor involvement, specifically involving a transition to bilateral stiffening (tonic phase) and subsequent convulsions (clonic phase).

Behavioral generalization was marked based on analysis of video-EEG recordings and occurred on average 89.0 ± 78.1 seconds after seizure onset and 72.0 ± 43.1 seconds before seizure end.

Focal Impaired Consciousness (FIC; formerly focal impaired awareness, FIA) seizures were identified when awareness was impaired at any point during the seizure, as indicated by a diminished ability to respond to verbal commands and/or amnesia for ictal events. Consciousness was assessed both behaviorally and retrospectively, including patient reports, and further characterized using dimensions from the Consciousness in Seizures Scale (CSS; Arthuis et al., 2009), specifically interaction with the examiner (CSS level 3) and recall of the seizure experience (CSS level 4). Focal Preserved Consciousness (FPC; formerly focal aware, FA) seizures were defined as those in which patients retained full awareness throughout the ictal period. These patients were able to interact appropriately with their environment and provided accurate retrospective reports of seizure events.

### Microelectrodes and intracranial data preprocessing

As a proxy for low-frequency local-field potentials from microelectrodes, raw 30 kHz micro-electrode data (0.25 µV . count^−1^) were down-sampled to 500 Hz, band-pass filtered 0.7–40 Hz (4^th^-order, zero-phase Butterworth), and shown with a 30 second baseline and the ictal segment, each plotted on a symmetric ± µV scale equal to the larger absolute excursion of the two epochs (Figure 1A).

**Figure 1.**
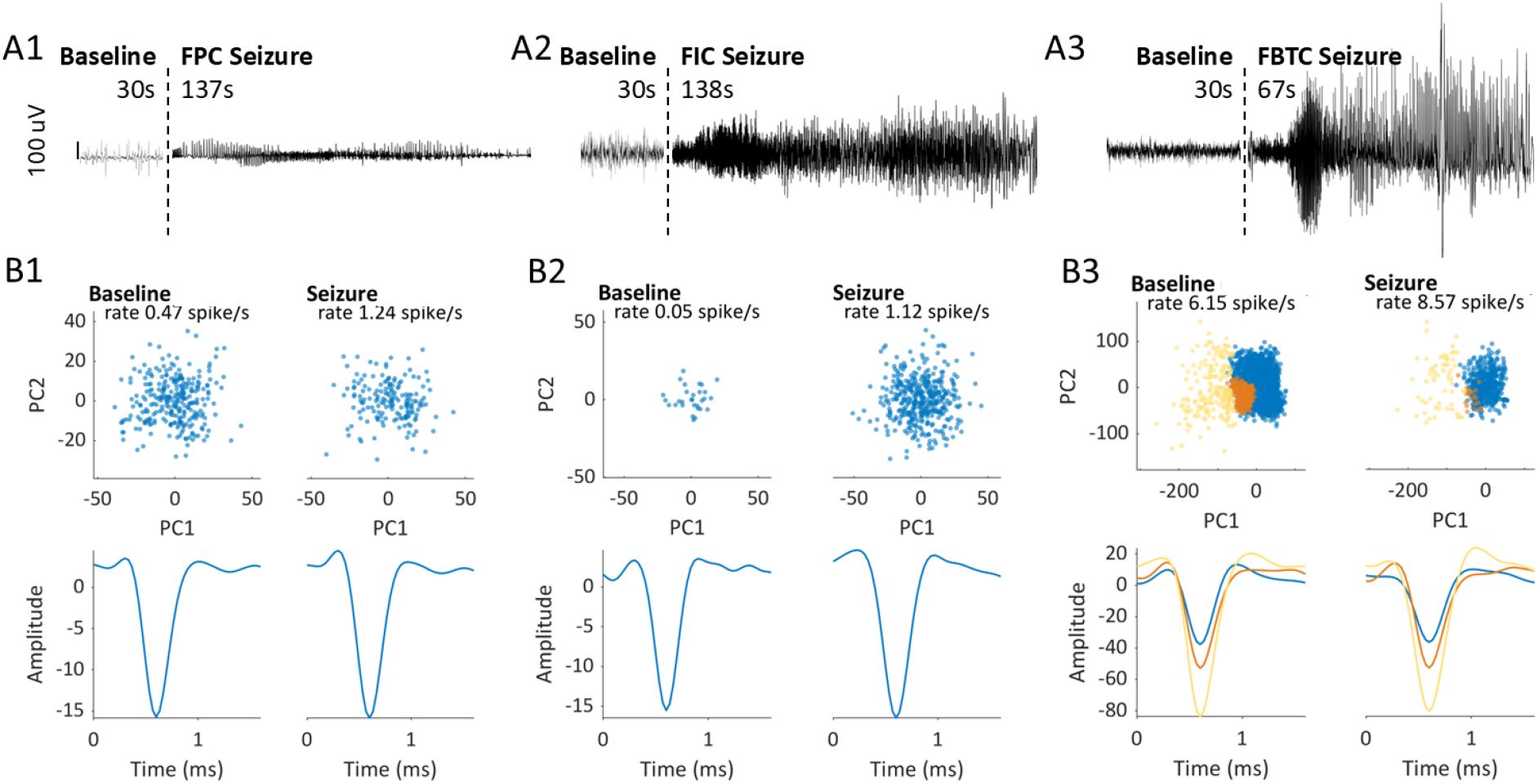
LFP microelectrode traces and spike features on seizure-onset zone (SOZ) channels from (A1) FPC, (A2) FIC, and (A3) FBTC seizures. Signals were down-sampled to 500 Hz and band-pass filtered 0.7– 40 Hz. For each contact, a 30-s interictal baseline (left) and the subsequent ictal segment (right) are shown on a common symmetric ±µV scale. (B1–B3) Spike features from the same seizures: PC1 vs. PC2 scatter plots for spikes from the 10-min preictal baseline and the ictal period. Spike sorting and PC computation were performed jointly over the full preictal + ictal window; panels are separated only for display. Overlaid traces are per-cluster mean waveform(s) for each period; note that all quantitative analyses pooled single-unit and multi-unit clusters and treated them as MUA.

For each seizure, a 10-minute pre-ictal baseline along with the entire ictal period was extracted for analysis. Microelectrode signals were band-pass filtered between 300–3000 Hz, and spike waveforms were detected as negative deflections exceeding 4.5 standard deviations from the mean (Hill et al., 2012; Quiroga et al., 2004). Spike sorting was performed on the window including both 10-minute pre-ictal and ictal periods, allowing for tracking unit changes from baseline to seizure. This approach favors stable, high–signal-to-noise units, as spikes that undergo distortion during local recruitment may fail detection or clustering. As a result, the analyses primarily reflect neurons that remain reliably sortable throughout the seizure.

Spike sorting was conducted semi-automatically using the UltraMegaSort2000 toolbox in MATLAB 2024 (MathWorks Inc., Natick, MA, USA), combining k-means clustering with visual inspection. Clusters visually identified as non-neuronal line-noise artifacts were excluded, and the remaining units, including both single units and multi-unit activity (MUA), were pooled and treated as MUA for all analyses.

For each microelectrode, MUA firing rates were calculated by binning spike times into 1-second non-overlapping windows, then averaging across contacts and normalizing to the 10-minute pre-ictal baseline. These firing rates were then analyzed in relation to seizure timing and anatomical location. Specifically, we compared firing rates between contacts located within (SOZ) and outside the seizure onset zone (non-SOZ). Firing rate comparisons were performed across the first and second halves of FPC and FIC seizures, or across pre- and post-generalization periods for focal to bilateral tonic-clonic (FBTC) seizures.

### Macroelectrode processing

Macroelectrode recordings were analyzed by extracting each seizure along with a 5-minute pre-ictal period, which served as the baseline for normalization. Signals were preprocessed using custom scripts incorporating EEGLAB routines (Delorme & Makeig, 2004) in MATLAB 2024 (MathWorks Inc., Natick, MA, USA). The raw signal was resampled to 400 Hz with an anti-aliasing filter and band-pass filtered from 0.5 to 199 Hz using a Hamming-windowed sinc FIR filter. Line noise was removed as necessary using the CleanLine algorithm, primarily at 60, 120, and 180 Hz. Channels and epochs with significant artifacts, such as motion artifacts, were excluded based on visual inspection. The power spectral density for each contact was computed using the ‘pwelch’ function, with a 10-second window and a 1-second overlap. Power spectrum values were then averaged across high-gamma (HG; 80–150 Hz) and delta (slow-wave activity, SWA; 1–4 Hz; see Supplementary material) bands. PLHG was computed as Hilbert transform of the high – (80-150 Hz) and low-pass (4-30 Hz) filtered signals for each electrode. The PLHG index was then computed within non-overlapping 1 second sliding windows as in Juan, Gorska, Kozma et al., 2023. All power markers were compared across FPC, FIC, and FBTC seizures. To enable comparison between micro and macroelectrodes, analyses were conducted on macroelectrode contacts positioned within and outside the SOZ, limited to areas extending up to 1.5 cm around the given microelectrode.

### Statistics

Group-level comparisons of neural activity were conducted separately for microelectrode firing rates and macroelectrode power measures (high-gamma, PLHG, and delta [see Supplementary Materials]) using linear mixed-effects (LME) models in R (R Core Team, 2015), implemented with the lme4 and lmerTest packages. For both datasets, the dependent variable (either log-transformed multi-unit firing rate or normalized power value) was modeled as a function of seizure type and ictal period, combined into a single categorical variable (seizure_period, e.g., “FIC-1st”, “FIC-2nd”, “PreG-FBTC”, “PostG-FBTC”) and electrode location relative to the seizure onset zone (SOZ vs. non-SOZ) was also included as a fixed effect, along with their interaction (seizure_period * SOZ). Subject and seizure identifiers were modeled as nested random intercepts to account for repeated measures. Estimated marginal means were obtained using emmeans (Lenth, 2021), with pairwise contrasts tested with false discovery rate (FDR) method. Only significant contrasts were visualized. Model assumptions were verified through residual diagnostics; minor deviations from normality and homoscedasticity were present for microelectrode models, but acceptable for LLM modeling. Models were fitted using Restricted Maximum Likelihood (REML), and all analyses and visualizations were conducted in R version 4.4.0.

## 3. RESULTS

### Multi-unit activity (MUA) is reliably trackable throughout seizures

We analyzed MUA from 32 seizures with various behavioral types (9 FPC, 17 FIC, 6 FBTC; see Methods) revealing stable clusters across pre-ictal and ictal epochs (Fig. 1). On SOZ channels, the micro-LFP showed a clear onset-related low-frequency transition on the same scale as baseline across seizure types (Fig. 1A). This evolution co-occurred with stable spike clusters (Fig. 1B), indicating that single unit activity recorded with Behnke–Fried electrodes can be tracked throughout seizures. Across all 895 units analyzed, waveform metrics were highly stable: ictal amplitudes closely matched baseline (median ictal/pre ≈ 1.00), and widths were centered near 1 overall, with slight broadening in SOZ for FBTC (median 1.08) and FIC (median 1.10).

### Ictal firing segregates by seizure type and SOZ proximity

Binned firing rates differed systematically across seizure types and between contacts within the seizure-onset zone (SOZ) versus outside (non-SOZ). For representative FPC seizure (see Fig. 2A), the SOZ contact exhibited an onset-locked rise that returned toward baseline across the ictal period, whereas the non-SOZ contact remained at baseline throughout. For representative FIC seizure (see Fig. 2B), the SOZ contact rose at onset and then attenuated later in the seizure, while firing outside the SOZ displayed a delayed increase. For representative FBTC seizure (see Fig. 2C), the SOZ contact rose at onset and remained elevated without an additional increase at generalization, whereas the non-SOZ contact displayed a sustained late increase time-locked to behavioral generalization.

**Figure 2.**
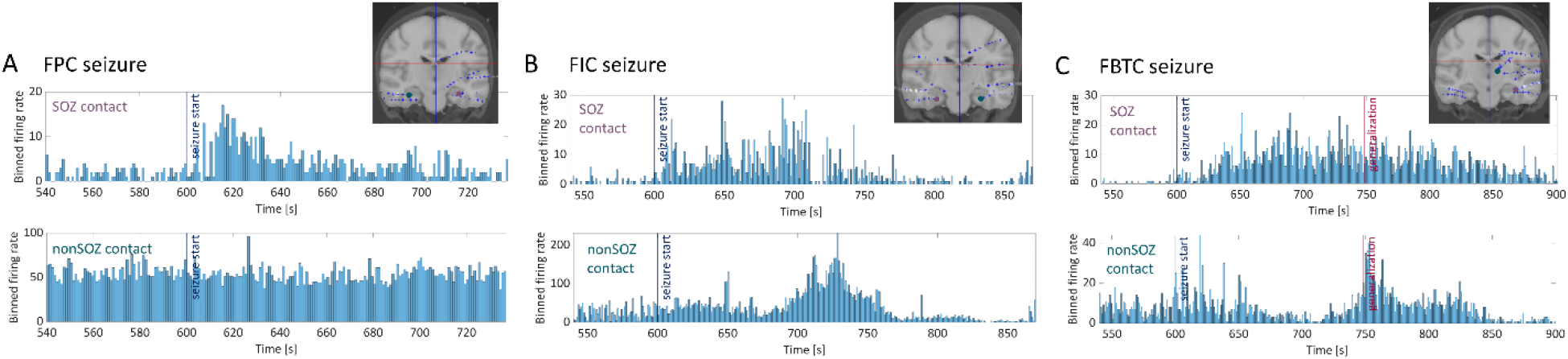
Example binned firing rates from single microelectrode contacts within the SOZ (top row; panel label in purple “SOZ contact”) and outside the SOZ (bottom row; panel label in teal “nonSOZ contact”) for representative seizures of three types: (A) FPC, (B) FIC, and (C) FBTC. Vertical lines indicate seizure onset (dark blue) and—only in FBTC—generalization (red). Insets show the SOZ and nonSOZ microelectrode contact locations on a common MRI from electrode reconstructions (coronal view).

Group-level microelectrode analysis included 6 FPC, 5 FIC, and 3 FBTC microelectrode sets in the SOZ, and 15 FPC, 33 FIC, and 8 FBTC sets outside the SOZ (Table 1 includes respective contact numbers). Group statistics (linear mixed-effects with FDR correction; Fig. 3) corroborated the single-contact patterns (Fig. 2). Within the SOZ (Fig. 3A), firing rate for FPC seizures increased in the first half (p < 0.05, ∼1-fold) and returned toward baseline in the second half (NS, ∼0.5-fold). Firing rate for FIC seizures was robustly elevated in both halves (p < 0.001 each; ∼2.4 and ∼1.6-fold increase, respectively) with attenuation in the second half (p < 0.01 for between halves contrast). Finally, firing rate for FBTC seizures was already elevated before generalization (p < 0.001, ∼1.6-fold) and remained so after it (p < 0.001, ∼1.5-fold), with no additional increase at generalization.

**Figure 3.**
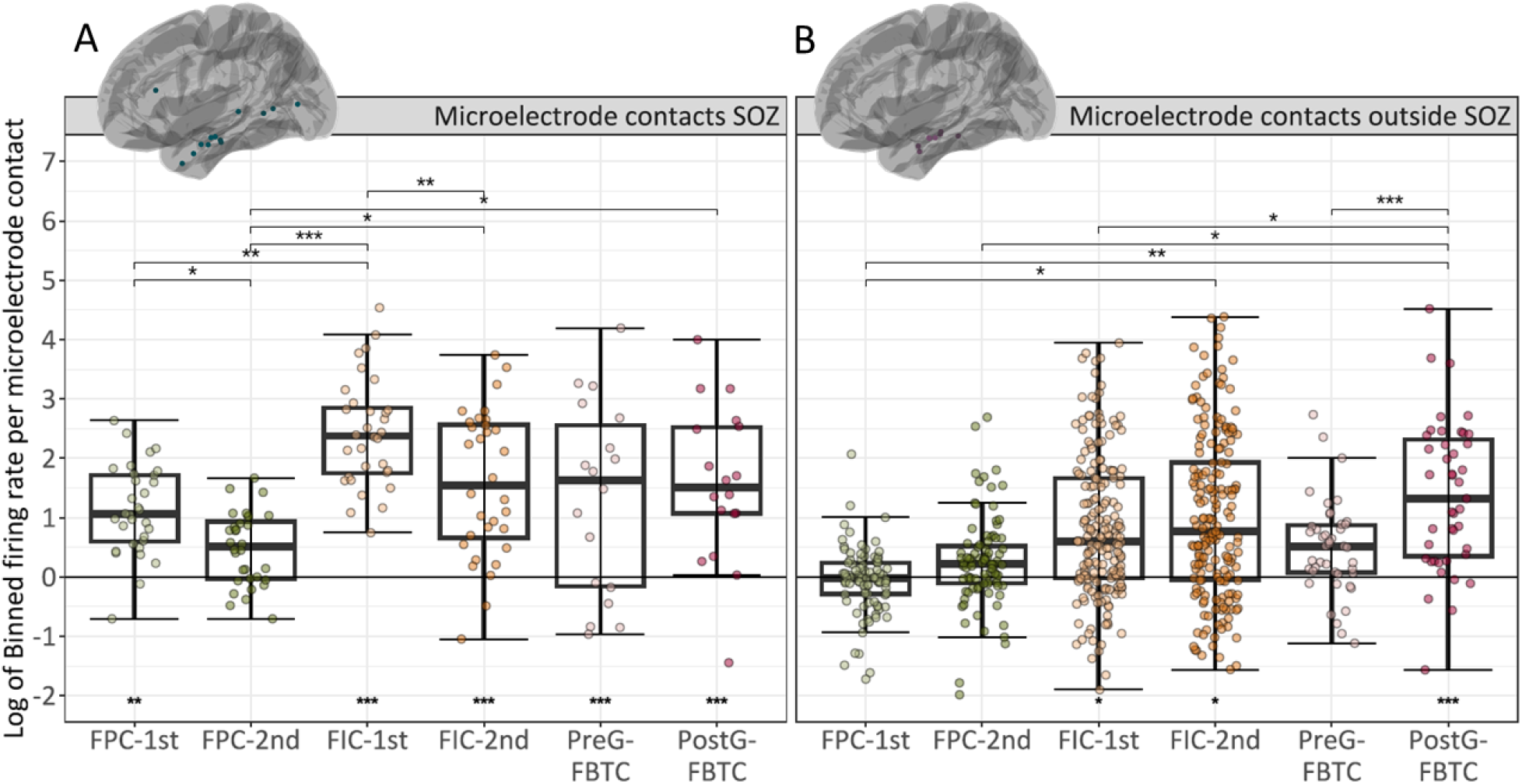
Log-binned firing rate per microelectrode contact for the first and second halves of FPC (light/dark green) and FIC (light/dark orange), and for pre- (pink) and post-generalization (red) periods of FBTC for contacts outside (A) and in SOZ (B). In each box plot, the bold horizontal line marks the median, the box spans the interquartile range (Q1–Q3), whiskers extend to 1.5×IQR, and points are individual contacts (values beyond the whiskers are outliers). Significance vs baseline (horizontal line at 0; log scale) is shown beneath each box, and pairwise differences between seizure periods are shown with bracket stars above—both from a linear mixed-effects model with FDR correction. (A) Outside-SOZ (teal) and (B) SOZ (purple) contacts are displayed on a left-lateral MNI surface above each plot. Stars indicate significance levels: p<0.05 (*), p<0.01 (**), p<0.001 (***); no star = not significant.

For microelectrodes outside the SOZ (Fig. 3B), firing rate for FPC did not differ from baseline in either half (both NS), FIC showed significant elevations in both halves (p < 0.05 each, ∼0.6-fold and ∼0.8 fold, respectively), and FBTC seizures were not significantly elevated before generalization (NS, ∼0.5-fold) but rose sharply after (p < 0.001, ∼1.3-fold).

### Macroelectrode high-gamma and PLHG track seizure-type–specific dynamics

Within the SOZ (Fig. 4B1), high-gamma (HG) was above baseline in FPC during the first half (p<0.05, ∼0.5 -fold) and returned towards baseline in the second half (NS; no difference between halves). In FIC, HG was elevated above baseline in both halves (p<0.05 each, ∼1-fold and ∼0.6-fold, respectively) without a significant half-to-half change. In FBTC, HG rose sharply above baseline during pre-generalization (p <0.05, ∼3-fold) and increased further post-generalization (p <0.01, ∼4.2-fold), with post-generalization values higher than pre-generalization (p <0.001), FPC and FIC (all p < 0.01). This contrasted with the firing rate patterns, where FBTC activity plateaued after generalization, whereas HG continued to increase.

**Figure 4.**
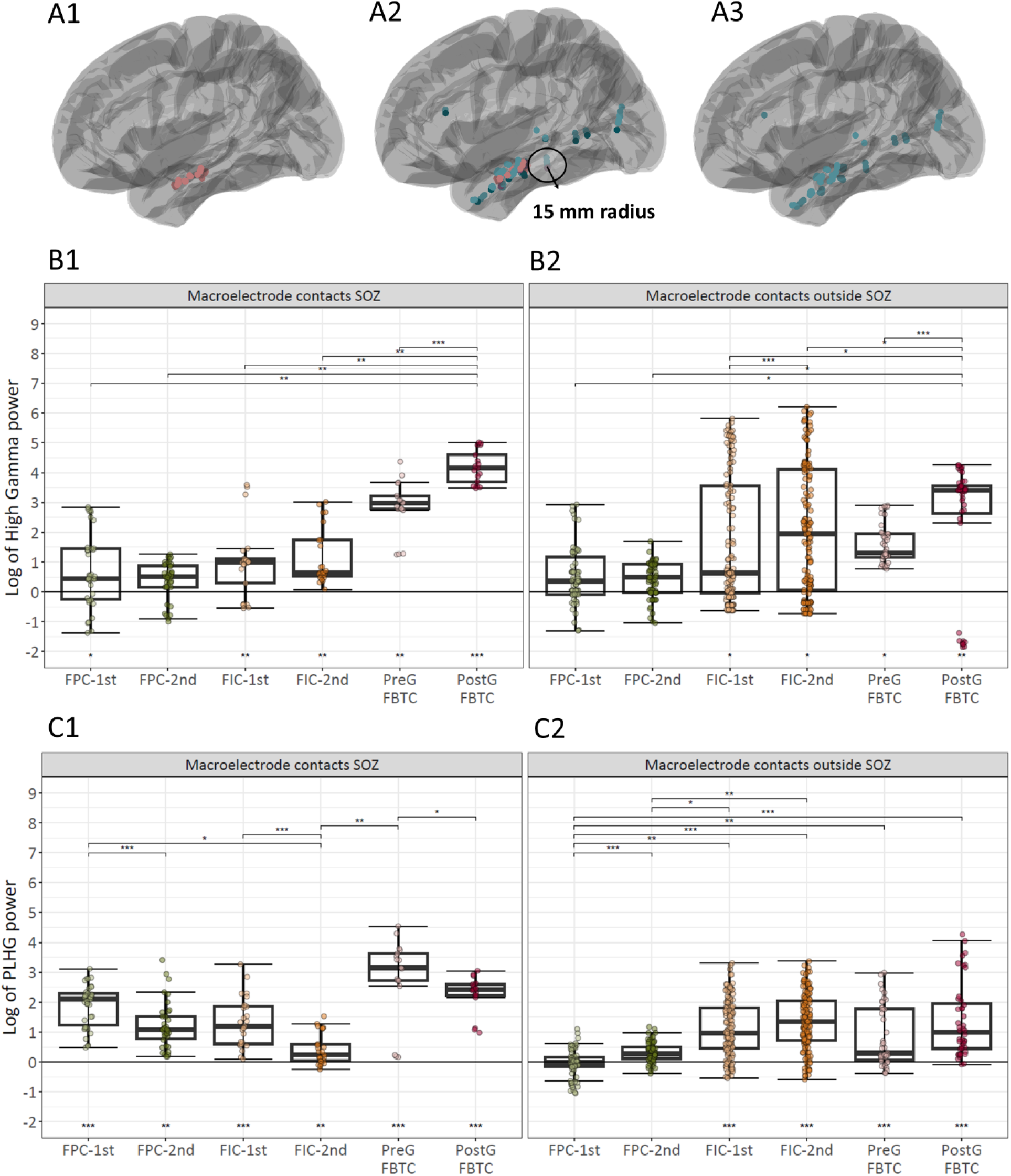
High-gamma, and phase-locked high-gamma (PLHG) power for the first and second halves of FPC (light/dark green) and FIC (light/dark orange), and for pre-(pink) and post-generalization (red) periods of FBTC computed on macroelectrodes surrounding microelectrodes localized either inside (BC1) or outside SOZ (BC2). In each box plot, the bold horizontal line marks the median, the box spans the interquartile range (Q1–Q3), whiskers extend to 1.5×IQR, and points are individual macroelectrode contacts (values beyond the whiskers are outliers). Macroelectrodes were selected as those within a 15-mm radius around each microelectrode (A2) and categorized as SOZ (A1; dark pink) and non-SOZ (A3; light teal) according to the microelectrode they surrounded. Significance versus baseline (horizontal line at 0; log scale) is shown beneath each box, and pairwise differences between seizure periods are shown with bracket stars above—both from a linear mixed-effects model with FDR correction. Stars indicate significance levels: p<0.05 (*), p<0.01 (**), p<0.001 (***); no star = not significant.

Within the SOZ (Fig. 4C1) PLHG rose above baseline at onset in all seizure types (first-half FPC 2.2-fold, FIC ∼1.2-fold, and pre-generalization FBTC ∼3.2-fold; all p < 0.001) and then declined over time: FPC and FIC fell from first to second half (to 1.1-fold and ∼0.2-fold respectively; between-half p < 0.001), and FBTC decreased from pre-to post-generalization (p < 0.05, ∼2.4-fold). This differed from firing rate in FBTC, which remained unchanged across generalization, whereas PLHG declined from pre-to post-generalization. Outside the SOZ (Fig. 4C2) PLHG remained at baseline in FPC across both halves (NS; 0 and ∼0.3-fold). In FIC, PLHG was robustly elevated in both halves (p<0.001 vs baseline, ∼0.9-fold and ∼1.3-fold, respectively with p < 0.01 between halves). In FBTC, PLHG was significantly elevated pre-generalization (p <0.001, ∼0.3-fold) and increased further post-generalization (p<0.001, ∼0.9-fold), with post-generalization values exceeding pre-generalization (p <0.001). Thus, outside the SOZ, the PLHG time course mirrored the firing rate pattern in FPC and FIC, whereas in FBTC it was already elevated pre-generalization, while firing rate increased only post-generalization.

Outside the SOZ (Fig. 4B2), HG power in FPC remained at baseline across both seizure halves (both NS; ∼0.3 and ∼0.5-fold, respectively). In FIC, HG was heterogeneous but consistently elevated relative to baseline, approximating a ∼0.5-fold increase in the first half and ∼2-fold increase in the second (each p < 0.05), with the second half exceeding the first (p < 0.001). In FBTC, HG rose at seizure onset (∼1.3-fold pre-generalization; p < 0.05) and strongly increased after generalization (∼3.9-fold; p < 0.05), with post-generalization values exceeding pre-generalization FBTC (p < 0.001) and both FIC and FPC halves (p < 0.05). In FBTC, the outside-SOZ increase in HG was slightly larger than matched change in microelectrode firing, which suggests widespread synchronous population activity above local neuronal recruitment.

## 4. DISCUSSION

This study demonstrated the feasibility of using Behnke–Fried microelectrodes to quantify multi-unit activity (MUA) through seizures and revealed distinct firing patterns inside versus outside the seizure-onset zone (SOZ), depending on the severity of seizures (FPC, FIC, FBTC). We found that within the SOZ, firing increased at onset for all seizure types and then attenuated in focal preserved consciousness (FPC) and focal impaired consciousness (FIC) seizures, versus plateaued in focal to bilateral tonic-clonic (FBTC) seizures. Outside SOZ, microelectrodes firing rate remained near baseline in FPC, rose gradually in FIC seizures, and displayed late increase that coincided with behavioral generalization in FBTC seizures. Macroelectrode markers around the same micro sites showed corresponding dynamics: in SOZ HG increases before generalization and rose further after generalization, while PLHG declined post-generalization even during FBTC seizures. Outside the SOZ, both HG and PLHG progressively increase over the course of FIC and FBTC seizures. Together, these micro-macro signatures suggest a practical way to differentiate SOZ, especially in FBTC, using the combination of early firing sustained through the seizure, accompanied by rising HG but falling PLHG at generalization.

In this work, using simultaneous microelectrode and macroelectrode recordings with hybrid Behne-Fried electrodes, we provided evidence that neuronal firing patterns differ systematically across behaviorally classified seizure types and by location (SOZ vs non-SOZ). Overall, we confirmed our recent work using Utah arrays, which showed a variable pattern of multi-unit firing outside SOZ before generalization, followed by a sustained, consistent increase after generalization, occurring alongside PLHG increases (Juan, Gorska, Kozma et al., 2023). We also examined FPC and FIC seizures, showing that firing rates, HG and PLHG outside SOZ did not change from baseline for FPC, but revealed stable firing rate and HG/PLHG increase from first to second half in FIC. Overall, outside-SOZ findings differentiate seizures with preserved versus impaired consciousness, with the latter revealing elevated firing, in line with increasing HG reflecting ictal activation in actively seizing areas (Eissa et al., 2016; Jirsch et al., 2006), and with increasing PLHG indexing locally synchronized, recruited cortex (Schevon et al., 2012; Weiss et al., 2013; Smith et al., 2016).

The firing rate effects we observed outside SOZ aligns with previous work suggesting that the narrow band of multi-unit firing migrates to other territories (Merricks et al., 2015) which would be reflected in stable elevated firing in FIC and pre-to post-generalization in FBTC seizures. These results are in line with observed multiple channels being recruited during FIC and significantly more and much more asynchronously during FBTC seizures (Juan, Gorska, Kozma et al., 2023). This pattern reinforces the possibility that a third driver amplifies cortical spread of activity beyond onset during generalization.

In the current work, we extended the analysis to SOZ firing rate dynamics and found that firing increased at onset for all focal seizure types investigated and then attenuated (FPC, FIC) or plateaued (FBTC). The observed pattern of onset peak with later attenuation matches the presence of an ictal wavefront driven by transient excitatory recruitment followed by a later recovery of postsynaptic inhibition during focal seizures (Smith et al., 2016; Merricks et al., 2021). Interestingly, in case of FBTC seizures we observed a plateau in firing instead of declining post-generalization which might be caused by possible involvement of additional driver from subcortical systems (Blumenfeld et al., 2009) given broad topography of an effect. This result needs to be confirmed on the larger sample.

The observed increase in firing followed by decline or continuation in SOZ corresponded with overall increases in HG in surrounding macroelectrodes, clearest for FBTC with further rise post-generalization. This HG increase at generalization confirms our previous findings (Juan, Gorska, Kozma et al., 2023), and - given that our macroelectrode coverage mainly includes limbic structures such as amygdala and hippocampus (Fig. 3A3) - it is compatible with engagement of deep limbic networks. Classic lesion and stimulation studies show that activating the brainstem reticular formation can rapidly drive cortical activation (Moruzzi & Magoun, 1949), elicit tonic components (Browning, 1985) and bilateral convulsions in generalized seizures (Gastaut et al., 1958). Recent Intracranial evidence suggests early, strong ictal recruitment of thalamus – specifically pulvinar (Pizzo et al., 2021) – with the anterior nucleus showing ictal LFP/high-gamma that scales with seizure severity (Singh et al., 2024) and the centromedian nucleus exhibiting robust ictal and post-ictal rhythmic activity (Arredondo et al., 2024).

Consistent with its coordinating role, thalamo-cortical coupling strengthens both seizure onset and termination (Soulier et al., 2023; Evangelista et al., 2015). Among subcortical candidates, the zona incerta is a plausible candidate that links reticular–thalamic circuits: it forms a central hub linking brainstem and thalamus via dense bilateral connections to intralaminar and higher order nuclei (Wang et al., 2020; Power & Mitrofanis, 2001), and it is the most sensitive locus for cholinergic induction of generalized seizures in basal forebrain (Brudzynski et al., 1995). This projection pattern suggest that subcortical engagement could amplify and bi-lateralize cortical activation – compatible with HG strengthening that we observed post-generalization.

In contrast to total HG power, PLHG confirmed our previous findings outside SOZ (Juan, Gorska, Kozma et al., 2023), while instead of increase, we observed decrease is SOZ during late ictal epochs (second halves, post-generalization). This is in line with previous work indicating that PLHG indexes recruited cortex (Schevon et al., 2012; Smith et al., 2016; Weiss et al., 2013), localizing ictal core with clinical utility (Weiss et al., 2015) and can be contrasted with non-recruited sites, where HG remains elevated (Agopyan-Miu et al., 2023). In contrast HG was shown not only to scale with population activation in the ictal core (Eissa et al., 2016), but also to spread outside SOZ being dependent on the global state (Bagshaw et al., 2009) and specifically increasing with FBTC generalization with strongest increases from deep sources (Juan, Górska, Kozma et al., 2023). Taken together, early firing and PLHG increase observed in our data in SOZ are in line with local recruitment, and later HG increase with attenuating PLHG could mark propagation with weaker synchrony, while post-generalization firing plateau suggests sustained drive from additional sources. Importantly, concomitant outside-SOZ increases in firing and HG/PLHG support additional drivers that amplify spread of seizures with increased severity (FIC, FBTC) and impaired consciousness.

This study has several limitations. First, the sample size was modest with relatively few FBTC seizures and only a subset of seizures sampled within the SOZ. While this allowed for initial group-level comparisons, larger cohorts will be needed to confirm seizure-type–specific differences. Second, anatomical coverage and SOZ location was limbic predominant, and only a fraction of microelectrodes was located within the clinically defined SOZ. Moreover, hybrid Behnke–Fried electrodes place the microelectrode brush only at the lead tip and space compared with Utah arrays (Chari et al., 2020), which all together limited our ability to capture neuronal populations comprehensively within each anatomical region. We did not record subcortical structures, preventing a direct test of thalamic or brainstem contributions that have been implicated in FBTC generalization (Englot et al., 2010; Doss et al., 2024).Our rigorous spike-sorting across extended preictal and full ictal epochs, necessarily favored stable high signal-to-noise units, likely underestimating recruitment that degrades spike shape or cluster separation (Merricks et al., 2015; Schevon et al., 2012), biasing against the most strongly recruited cells. Future work will collect a broader sample with more diverse anatomical coverage to confirm these findings.

## CONCLUSIONS

This work demonstrates feasibility of using hybrid Behne-Fried electrodes to track multi-unit activity (MUA) through the ictal period in seizures with preserved and impaired consciousness. We identify location-specific dynamics: within the SOZ, firing rises at onset and then attenuates (FPC, FIC) or plateaus (FBTC); outside the SOZ, firing remains near baseline in FPC, rises gradually in FIC, and increases sharply at FBTC generalization. In co-localized macro-electrodes, PLHG behaves as a recruitment marker, increasing early and then attenuating, while HG reflects broader activation, building outside the SOZ and especially after FBTC generalization. Together, these micro-macro patterns may provide new SOZ biomarkers and key ground for further mechanistic and clinical studies.

## Data Availability

All data produced in the present study are available upon reasonable request to the authors.

## Acknowledgement

The authors thank Rosario Ciciento, Liam Tabor, Marek Solvik, Jack Vogel, Cameron Brace, Alec Adamov, Hannah Holmes, and Camile Mejia. This work was supported by NIH (grant number 1K23NS112473), and Tiny Blue Dot foundation.

## Contributions

**Conceptualization:** Melanie Boly, **Data curation:** Urszula Gorska-Klimowska, Beril Mat **Formal analysis:** Urszula Gorska-Klimowska, Beril Mat, Maximilian Grobbelaar, Elena Monai, Felipe B. de Paiva, Cynthia Papantonatos, **Funding acquisition:** Melanie Boly, Giulio Tononi, **Investigation:** Brinda Sevak, Mariel Kalkach Aparicio, Dillon Scott, Csaba Kozma, Beril Mat, Cynthia Papantonatos, Aaron Suminski, Aaron F. Struck, Melanie Boly, Wendell Lake, **Methodology:** Urszula Gorska-Klimowska, Colin Denis, Melanie Boly, **Project administration:** Melanie Boly, Urszula Gorska-Klimowska, Beril Mat, **Resources:** Melanie Boly, Giulio Tononi, Aaron F. Struck, Aaron Suminski, **Software:** Urszula Gorska-Klimowska, Colin Denis, Felipe B. de Paiva, Csaba Kozma, **Supervision:** Melanie Boly, Urszula Gorska-Klimowska, **Validation:** Urszula Gorska-Klimowska, **Visualization:** Urszula Gorska-Klimowska, **Writing – original draft:** Urszula Gorska-Klimowska, **Writing – review & editing:** Urszula Gorska-Klimowska, Melanie Boly, Aaron Suminski.

## Conflict of interest

None of the authors have any conflict of interest to disclose.

We confirm that we have read the Journal’s position on issues involved in ethical publication and affirm that this report is consistent with those guidelines.

## Data and code availability

De-identified data underlying this study are available from the corresponding author upon reasonable request, subject to IRB/ethics approval and a data use agreement. All analysis scripts used will be also shared on request.

