## Supplementary materials for "Multi-unit activity and high-gamma patterns dissociate in the seizure onset zone during focal to bilateral tonic-clonic seizures"

To assess consistency of unit-level recruitment, we modeled spikes as Poisson events and flagged significant firing-rate changes when the observed seizure spikes differed from the expected count by $|z| > 3$ of a unit-specific Poisson distribution (following Merricks et al., 2015). We then aggregated the proportion of significantly modulated units by seizure type, SOZ status, and ictal period. This analysis complements population-level models by testing whether observed firing-rate dynamics reflect coordinated recruitment across units, rather than only mean-rate shifts.

In FPC seizures, SOZ units showed high rates of increase (88% in the first half and 75% second half of the seizure, respectively) while non-SOZ changes were modest (26%, and 52%). In FIC seizures, 100% of SOZ units increased in both halves versus 63–83% outside SOZ. In FBTC seizures, SOZ modulation rose from 81% pre-generalization to 100% post-generalization, whereas non-SOZ shifted from 100% pre-generalization to 85% post-generalization.

Overall, the unit-level Poisson test confirmed group level firing-rate patterns, with only divergence for non-SOZ FBTC contacts revealing pre- to post-generalization increase whereas the Poisson analysis showed a drop in the fraction of significantly modulated units.

Delta power analysis was described in the Methods section.

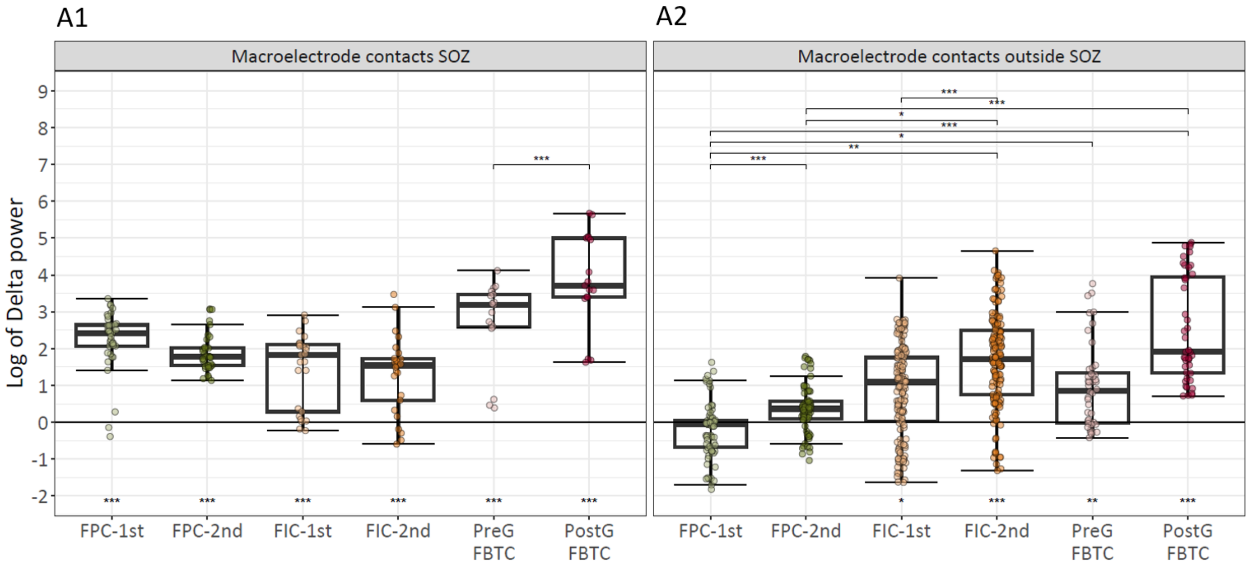


**Supplementary Figure 1.** Delta (1–4 Hz) power for the first and second halves of FPC (light/dark green) and FIC (light/dark orange), and for pre- (pink) and post-generalization (red) periods of FBTC, computed on macroelectrodes surrounding microelectrodes localized either inside SOZ (A1) or outside (A2). In each box plot, the bold horizontal line marks the median, the box spans the interquartile range (Q1–Q3), whiskers extend to 1.5×IQR, and points are individual contacts (values beyond the whiskers are outliers). Significance versus baseline (horizontal line at 0; log scale) is shown beneath each box, and pairwise differences between seizure periods are shown with bracket stars—both from linear mixed-effects models with FDR correction. Stars indicate $p<0.05$ (*), $p<0.01$ (**), $p<0.001$ (***); no star = not significant.

Within the SOZ, delta (1–4 Hz) power was elevated relative to baseline in all periods and seizure types (all $p < 0.001$). Across time, only FBTC showed a reliable step-up from pre- to post-generalization ($p< 0.001$; from ~3-fold to ~3.6-fold), while FPC and FIC remained stable with slight tendency to decrease in the second half (~2.4-fold to ~1.7-fold for FPC, and ~1.7-fold to ~1.5-fold FIC).

Outside the SOZ, delta power remained at baseline in FPC across halves (both NS, 0 and ~0.3-fold). In FIC and FBTC it followed a clear two-step course: a modest rise early (first half FIC ~1-fold, $p < 0.05$ vs baseline and pre-generalization FBTC ~0.8-fold, $p < 0.01$ vs baseline) that became more pronounced later (second half FIC ~1.7-fold and post-generalization FBTC ~1.9-fold; $p < 0.001$ vs baseline).

Taken together, slow-wave activity (delta power; 1–4 Hz) acted as a complementary, state-dependent index of ictal activity (Doss et al., 2024): it was elevated within SOZ with an additional post-generalization increase, and it rose outside SOZ in FIC and FBTC, with additional increase in second half and post-generalization. Mechanistically, SWA reflects cortical bistability: hyperpolarized down-states reduce information integration and likely contribute to LOC (Sarasso et al., 2014; Tononi et al., 2016), consistent with widespread sleep-like cortical rhythms during seizure-induced LOC (Englot et al., 2010). Accordingly, SWA might be related to propagation associated with impaired consciousness in FIC and FBTC, complementing the main findings.
